# Diagnostic Accuracy of the Abbot BinaxNOW COVID-19 Antigen Card Test, Puerto Rico

**DOI:** 10.1101/2023.12.14.23299967

**Authors:** Zachary J. Madewell, Chelsea G. Major, Nathan Graff, Cameron Adams, Dania M. Rodriguez, Tatiana Morales, Nicole A. Medina Lopes, Rafael Tosado, Liliana Sánchez-González, Janice Perez-Padilla, Hannah R. Volkman, Jorge Bertran, Diego Sainz, Jorge Munoz-Jordan, Gilberto A. Santiago, Olga Lorenzi, Vanessa Rivera-Amill, Melissa A. Rolfes, Gabriela Paz-Bailey, Laura E. Adams, Joshua M. Wong, PRESCA Study Team

**Author notes:** **Corresponding author**: Zachary J. Madewell **Corresponding author Email:**.

## Abstract

**Background:** The COVID-19 pandemic underscored the need for rapid and accurate diagnostic tools. In August 2020, the Abbot BinaxNOW COVID-19 Antigen Card test became available as a timely and affordable alternative for SARS-CoV-2 molecular testing, but its performance may vary due to factors including timing and symptomatology. This study evaluates BinaxNOW diagnostic performance in diverse epidemiological contexts.

**Methods:** Using RT-PCR as reference, we assessed performance of the BinaxNOW COVID-19 test for SARS-CoV-2 detection in anterior nasal swabs from participants of two studies in Puerto Rico from December 2020 to May 2023. Test performance was assessed by days post symptom onset, collection strategy, vaccination status, symptomatology, repeated testing, and RT-PCR cycle threshold (Ct) values.

**Results:** BinaxNOW demonstrated an overall sensitivity of 84.1% and specificity of 98.8%. Sensitivity peaked within 1–6 days after symptom onset (93.2%) and was higher for symptomatic (86.3%) than asymptomatic (67.3%) participants. Sensitivity declined over the course of infection, dropping from 96.3% in the initial test to 48.4% in testing performed 7–14 days later. BinaxNOW showed 99.5% sensitivity in participants with low Ct values (≤25) but lower sensitivity (18.2%) for participants with higher Cts (36–40).

**Conclusions:** BinaxNOW demonstrated high sensitivity and specificity, particularly in early-stage infections and symptomatic participants. In situations where test sensitivity is crucial for clinical decision-making, nucleic acid amplification tests are preferred. These findings highlight the importance of considering clinical and epidemiological context when interpreting test results and emphasize the need for ongoing research to adapt testing strategies to emerging SARS-CoV-2 variants.

## Introduction

As of October 2023, the COVID-19 pandemic has led to 771 million confirmed cases of COVID- 19 and 7 million deaths globally, with Puerto Rico reporting almost 1.3 million COVID-19 cases and 6,000 associated deaths.^1^ Rapid identification of SARS-CoV-2 infection and subsequent measures to reduce transmission are central to an effective public health response to COVID-19.^2^ However, the broad spectrum of clinical manifestations of SARS-CoV-2 infection poses a challenge to the rapid identification of infections and the implementation of effective measures to reduce transmission.^2–4^ Concurrently, the pandemic prompted the development of novel therapies ^5–8^ that are designed to shorten COVID-19 symptom duration. Early identification of SARS-CoV-2 infection is crucial for the timely and appropriate administration of therapies, particularly for people at higher risk for severe disease. Many of the novel treatments developed during the pandemic require initiation within a specific window after symptom onset. However, the challenges posed by the broad spectrum of clinical manifestations, including initially asymptomatic and mild cases that can progress to severe disease, make early and accurate detection of SARS-CoV-2 infection essential for effective treatment and prevention strategies.

To identify infected individuals for isolation and appropriate medical therapy, rapid and accurate COVID-19 tests continue to play a crucial role, including those used in clinical and laboratory settings. Although RT-PCR-based testing is frequently available in clinical and laboratory settings for infection detection, its utility can be limited by the expertise required for proper sample management and reporting delays due to the time needed for transport and testing at laboratory facilities.^9,10^ In many communities, point-of-care rapid antigen tests were deployed to enhance the accessibility and efficiency of SARS-CoV- 2 infection detection. Among the available commercial lateral flow antigen tests, the BinaxNOW Antigen Card test has undergone particularly extensive evaluation, demonstrating consistent specificity (>97%) across multiple cohort studies.^11–15^ However, sensitivity estimates varied widely in different reports, with potential factors including timing of specimen collection, symptom presence, collection methodology, and viral replication levels, necessitating further validation.

In December 2020, BinaxNOW testing was introduced alongside RT-PCR testing for SARS-CoV-2 in a community cohort and two clinical surveillance sites in Puerto Rico. We evaluated how the performance of BinaxNOW varied by days post onset of symptoms, symptomatology, predominant SARS-CoV-2 variant, vaccination status, collection strategy, repeated tests, and RT-PCR cycle thresholds (Ct). This study leverages its large sample size, including specimens collected at various time points from a unique population in Puerto Rico, to provide a comprehensive evaluation of the BinaxNOW Antigen Card test’s performance, contributing to filling an information gap in the use of point-of-care rapid antigen tests for SARS-CoV-2 infection detection. Our findings contribute to a deeper understanding of the test’s efficacy and role in augmenting current diagnostic strategies.

## Methods

### Study Design and Data Collection

The data analyzed is derived from two observational studies in Puerto Rico: the Communities Organized to Prevent Arboviruses (COPA) study and the Sentinel Enhanced Dengue Surveillance System (SEDSS), both of which are conducted by the Ponce Health Sciences University (PHSU) and the US Centers for Disease Control and Prevention’s (CDC) Dengue Branch (DB).

COPA is a community-based cohort study established in Ponce, Puerto Rico, in 2018. Study enrollment and data collection activities are described elsewhere.^16–18^ Briefly, study activities include annual interviews and serum collection for arbovirus testing among approximately 3,800 participants. Beginning in April 2020, anterior nasal swabs for SARS-CoV-2 RT-PCR testing were collected from participants that reported experiencing COVID-like symptoms (i.e., fever, cough, sore throat, difficulty breathing, diarrhea, body pain, or loss of taste/smell) or within the last 7 days of their annual visits.

Additionally, an acute illness surveillance component was initiated via weekly text messages asking participants to report if they or a household member experienced COVID-like symptoms in the past 7 days. Symptomatic participants, as well as those with a prior positive lab test for SARS-CoV-2 in the last 7-21 days and their household contacts, were offered visits for anterior nasal swab collection for SARS-CoV-2 RT-PCR testing. Beginning in December 2020, concurrent collection of a second anterior nasal swab for testing by the BinaxNOW COVID-19 Antigen Card test was offered to all participants with a swab collected for SARS-CoV-2 RT-PCR testing. All nasal swabs were collected by study staff, and BinaxNOW testing was performed within one hour of collection at the study site. Our analyses include COPA participants who were tested for SARS-CoV-2 between December 2020 and May 2023 using both BinaxNOW and RT-PCR assays. COPA participants may have been tested multiple times in the study period, including during the same and separate illness or exposure events.

Established in May 2012, SEDSS is an active surveillance system that monitors acute febrile and respiratory illnesses in two emergency departments in Ponce, Puerto Rico. In 2018, an additional site was established in an emergency department in San Juan.^21–23^ Patients were eligible for enrollment if they demonstrated fever upon presentation or within the past week (oral temperature ≥38°C, axillary temperature ≥38.5°C), or cough/dyspnea within the last 14 days (with or without fever). Nasopharyngeal swabs collected at enrollment from participants in SEDSS were tested for SARS-CoV-2 using RT-PCR. Two collection approaches were employed for BinaxNOW testing in one of the two participating emergency departments: staff-collected and participant-collected (self-testing) anterior nasal swabs.

Participants underwent staff-collected, self-collected, or both staff- and self-collected anterior nasal swabbing concurrently. Participants were provided with clear and simple instructions for self-collection and testing, including applying drops to the test card, swabbing both nostrils, and following specific steps for test card handling.^25^ Our analyses included SEDSS participants in the San Juan or Ponce sites tested for SARS-CoV-2 between January and April 2021 using both BinaxNOW and RT-PCR assays.

For both COPA and SEDSS, the RT-PCR assays used included the CDC Real-Time Reverse Transcription PCR Panel for tests performed before December 2021 and the CDC Influenza SARS-CoV- 2 (Flu SC2) Multiplex Assay for tests performed December 2021 and later.^19,20^

### Statistical Analysis

We reported frequencies of demographic characteristics (age group, sex, ethnicity, race, and Hispanic/Latino), reported chronic medical conditions, COVID-19 vaccine doses, and number of RT-PCR/BinaxNOW tests among all COPA and SEDSS participants with one or more RT-PCR/BinaxNOW test result data available.

Using the SARS-CoV-2 RT-PCR result as our reference standard, we calculated measures of diagnostic accuracy of BinaxNOW tests including sensitivity, specificity, positive predictive value, negative predictive value, positive likelihood ratio, negative likelihood ratio, and the number needed to diagnose (NND) of BinaxNOW tests compared to RT-PCR tests. Definitions of these measures are given in Table S1. We calculated 95% confidence intervals (CI) for all measures. We used McNemar’s test to evaluate differences in proportions of discordant pairs (i.e., the differences between false positives and false negatives) between BinaxNOW and the reference standard, RT-PCR.^26^ It helps determine if one test is more likely to produce false positives or false negatives compared to the other. To assess discrimination, we calculated the area under the receiver operating characteristic curve (AUC-ROC).

AUC-ROC summarizes the trade-off between sensitivity and specificity, where an AUC of 1 indicates perfect discrimination, and 0.5 indicates no discrimination.

We evaluated the performance of BinaxNOW compared to RT-PCR overall across all participants as well as by days post symptom onset (0, 1–3, 4–6, 7+ days), symptom status (asymptomatic, symptomatic), collection strategy (staff-collected, self-collected), number of COVID-19 vaccine doses received prior to testing (0, 1, 2, 3 doses), primary SARS-CoV-2 variant (pre-Delta, Delta, Omicron) circulating at time of sample collection, and Ct values of positive RT-PCR tests (≤25, 26–30, 31–35, 36– 40). The classification of primary circulating SARS-CoV-2 variant was based on the time period from their earliest detection in Puerto Rico until the detection of a new major variant: pre-Delta (cases through May 31, 2021), Delta (June 1 to November 30, 2021), and Omicron (after December 1, 2021).^27^

For COPA participants with repeated tests, we evaluated BinaxNOW performance for their initial test as well as the repeated test 7–14 days after the initial test. We further stratified this analysis by participant symptom status for the initial and repeated tests. We performed a sensitivity analysis for the repeated tests by restricting to participants who had testing within 6 days of symptom onset to ensure the repeated test was not for a different infection. For repeated tests among COPA participants, tests separated by ≥90 days were considered as part of separate illness episodes, and tests within 7–14 days of each other were considered part of the same illness episode.^28^ The few COPA participant tests performed between 15–89 days of another test were excluded from the analysis. In SEDSS, when both self-collected and hospital staff-collected swabs were tested, all tests, including the RT-PCR test, were conducted on the same day and included in the analyses.

We fit cubic splines to further understand the relationships between sensitivity and specificity of BinaxNOW by days post onset of symptoms and total number of symptoms. All analyses were done using R software, version 4.3.1 (R Foundation for Statistical Computing, Vienna, Austria).

## Results

There were 1,207 total participants with results from paired BinaxNOW and RT-PCR tests: 943 (78.1%) from COPA and 264 (21.9%) from SEDSS (Table 1). The median age of all participants was 36 years (IQR: 17, 49), 57.4% were female, 99.7% were Hispanic/Latino, and 56.3% had reported past diagnosis with one or more chronic medical conditions. Of 799 COPA participants with available COVID-19 vaccine data, 92.5% had received at least two doses, whereas 5.8% remained unvaccinated. All SEDSS participants were unvaccinated and tested before vaccines became widely available in Puerto Rico. Among the 264 SEDSS participants, 58 (22.0%) underwent both staff-collected/tested and participant-collected/tested BinaxNOW tests, resulting in a total of 322 BinaxNOW tests. In COPA, there were 1,208 BinaxNOW tests from the 943 participants from December 2020 to May 2023. Of the 1,530 total tests from SEDSS and COPA, 404 (26.4%) were positive for SARS-CoV-2 on the BinaxNOW test and 465 (30.4%) were positive by RT-PCR.

**Table 1.** Demographic characteristics of participants from COPA and SEDSS, 2020– 2023.

|  | Overall<br>N = 1207 | COPA<br>N = 943 | SEDSS<br>N = 264 |
| --- | --- | --- | --- |
| **Age in years** (median [IQR]) | 36 [16, 49] | 36 [16, 47] | 36 [19, 58] |
| **Age group in years (%)** (N = 1207) |  |  |  |
| 0–10 | 149 (12.3) | 96 (10.2) | 53 (20.1) |
| 11–20 | 222 (18.4) | 208 (22.1) | 14 (5.3) |
| 21–30 | 146 (12.1) | 102 (10.8) | 44 (16.7) |
| 31–40 | 180 (14.9) | 142 (15.1) | 38 (14.4) |
| 41–50 | 263 (21.8) | 237 (25.1) | 26 (9.8) |
| 51+ | 247 (20.5) | 158 (16.8) | 89 (33.7) |
| **Sex (%)** (N = 1206) |  |  |  |
| Female | 692 (57.4) | 552 (58.6) | 140 (53.0) |
| Male | 514 (42.6) | 390 (41.4) | 124 (47.0) |
| **Hispanic/Latino (%)** (N = 1171) |  |  |  |
| Yes | 1168 (99.7) | 912 (100.0) | 256 (98.8) |
| No | 3 (0.3) | 0 (0) | 3 (1.2) |
| **Ethnicity (%)** (N = 1170) |  |  |  |
| Puerto Rican | 1152 (98.5) | 900 (98.8) | 252 (97.3) |
| Other | 18 (1.5) | 11 (1.2) | 7 (2.7) |
| **Race (%)** (N = 1118) |  |  |  |
| Black | 122 (10.9) | 93 (10.4) | 29 (12.9) |
| Mixed | 101 (9.0) | 85 (9.5) | 16 (7.1) |
| White | 849 (75.9) | 685 (76.7) | 164 (72.9) |
| Other | 46 (4.1) | 30 (3.4) | 16 (7.1) |
| **Chronic medical conditions (%)** (N = 1205) |  |  |  |
| Yes | 679 (56.3) | 531 (56.3) | 148 (56.5) |
| No | 526 (43.7) | 412 (43.7) | 114 (43.5) |
| **COVID-19 vaccine doses recorded on final visit (%)** (N = 1063) |  |  |  |
| 0 | 310 (29.2) | 46 (5.8) | 264 (100) |
| 1 | 14 (1.5) | 14 (1.8) | 0 (0) |
| 2 | 290 (30.5) | 290 (36.3) | 0 (0) |
| 3 | 435 (45.7) | 435 (54.4) | 0 (0) |
| 4 | 14 (1.5) | 14 (1.8) | 0 (0) |
| **Symptomatic during study (%)** (N = 1203) |  |  |  |
| Yes | 1030 (85.6) | 770 (81.7) | 260 (100) |
| No | 173 (14.4) | 173 (18.3) | 0 (0) |
| **Days from symptom onset to testing**<br>(median [IQR]) (N = 923) | 4 [2, 6] | 4 [3, 7] | 2 [1, 4] |
| **Number of RT-PCR/BinaxNOW tests (%)** (N = 1207) |  |  |  |
| 1 | 733 (60.7) | 527 (55.9) | 206 (78.0) |
| 2 | 318 (26.3) | 260 (27.6) | 58 (22.0)<sup>a</sup> |
| 3 | 94 (7.8) | 94 (10.0) | 0 (0.0) |
| ≥4 | 62 (5.1) | 62 (6.6)<sup>b</sup> | 0 (0.0) |

IQR: interquartile range; COPA: Communities Organized to Prevent Arboviruses; SEDSS: Sentinel Enhanced Dengue Surveillance System; RT-PCR: reverse transcription polymerase chain reaction.
<sup>a</sup> All repeat testing for SEDSS participants was performed on the same day with one swab collected by a healthcare provider and another self-collected swab.
<sup>b</sup> For repeated tests among COPA participants, tests separated by ≥90 days were considered as part of separate illness episodes, and tests within 7–14 days of each other were considered part of the same illness episode. COPA participant tests performed between 15–89 days of another test were excluded from the analysis.

Across all participants (n=1,530 paired tests), the overall sensitivity of BinaxNOW compared to RT-PCR was 84.1% (95% CI: 80.4%–87.3%), specificity was 98.8% (95% CI: 97.9%–99.3%), positive predictive value was 96.8% (95% CI: 94.6%–98.3%), and negative predictive value was 93.4% (95% CI: 91.8%–94.8%) (Table 2). We further examined the diagnostic performance at different time intervals following symptom onset. Sensitivities at 1–3 days post onset (92.1%) and 4–6 days post onset (94.2%) were significantly higher than at ≥7 days post onset (70.2%) (*p*<0.001). Specificity remained consistently above 98% across all days post-onset. The sensitivity of the BinaxNOW test peaked between 1 and 6 days post-onset and waned thereafter (Figure 1).

**Figure 1.**
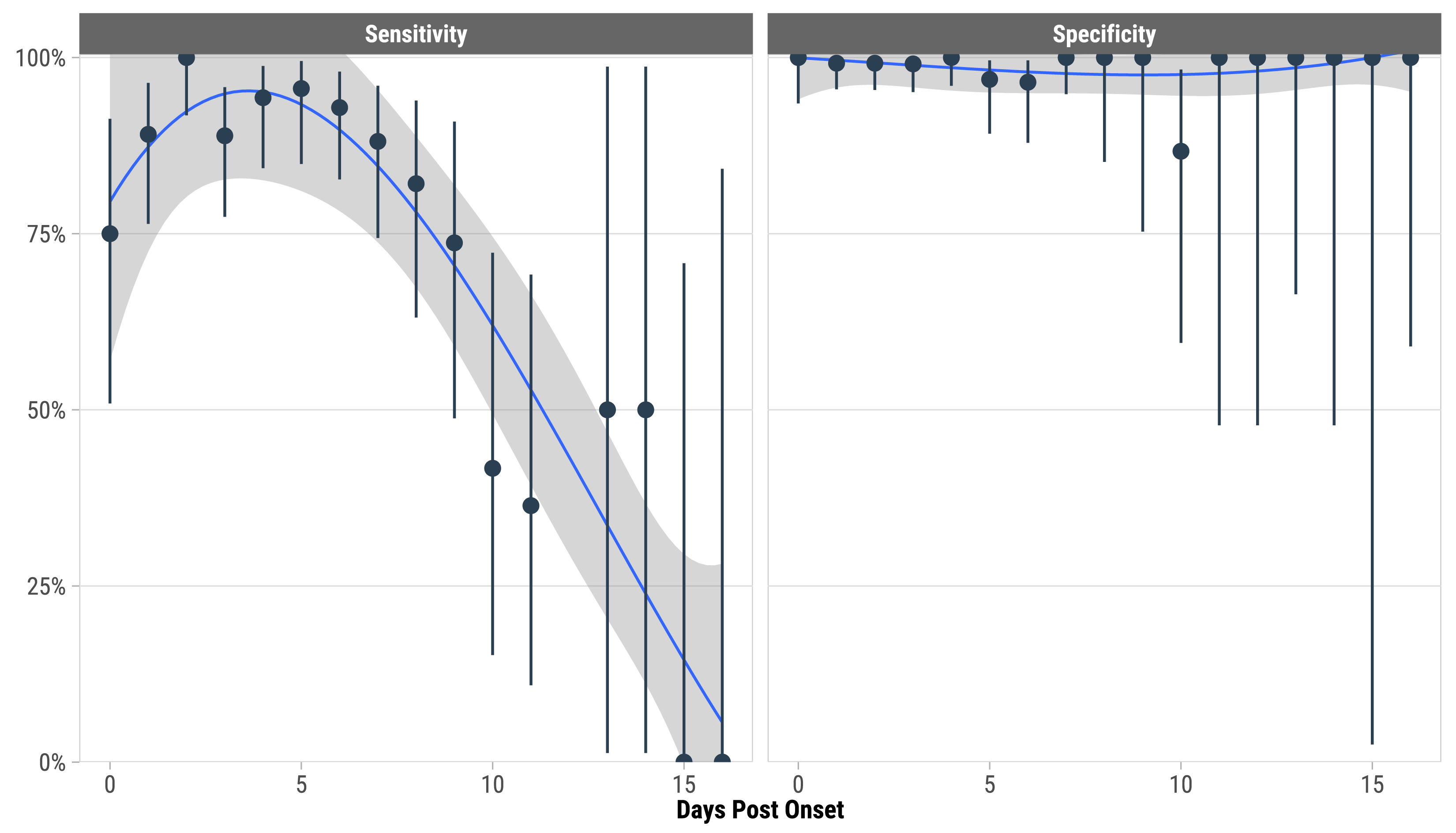
Sensitivity and specificity of BinaxNOW Antigen test compared to RT-PCR by days post onset of symptoms (N = 1181 paired tests from 921 participants with 0 to 16 days post onset). The blue line represents a cubic spline and grey bands indicate 95% confidence intervals of the model fit. Vertical bars are 95% confidence intervals of BinaxNOW sensitivity and specificity for each days-post- onset subgroup. There were 1181 tests of both BinaxNOW and RT-PCR.

**Table 2.**
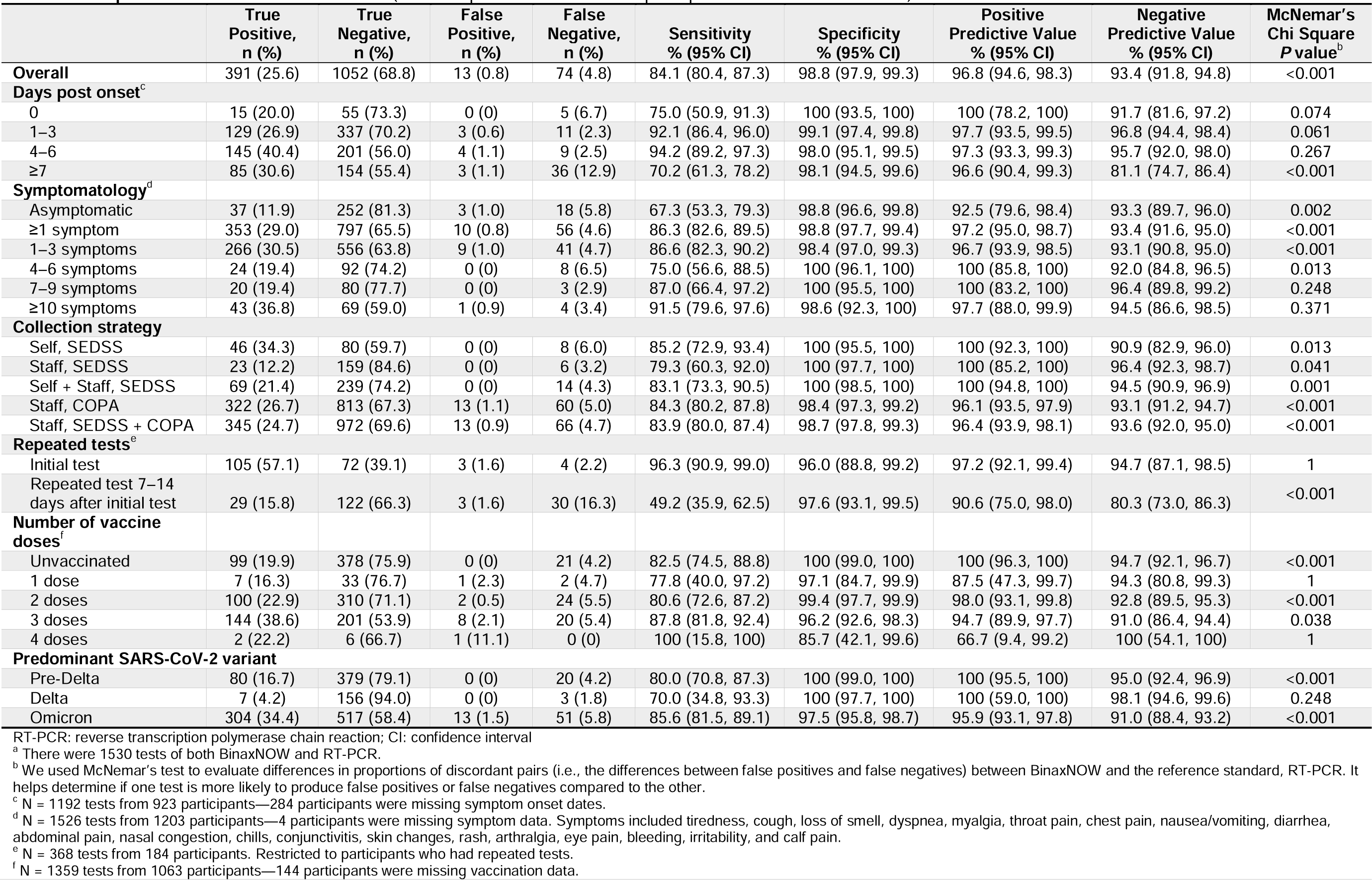
Comparison of BinaxNOW and RT-PCR (N = 1530 paired^a^ tests from 1207 participants unless stated otherwise).

|  | True Positive, n (%) | True Negative, n (%) | False Positive, n (%) | False Negative, n (%) | Sensitivity % (95% CI) | Specificity % (95% CI) | Positive Predictive Value % (95% CI) | Negative Predictive Value % (95% CI) | McNemar's Chi Square P value<sup>b</sup> |
| --- | --- | --- | --- | --- | --- | --- | --- | --- | --- |
| **Overall** | 391 (25.6) | 1052 (68.8) | 13 (0.8) | 74 (4.8) | 84.1 (80.4, 87.3) | 98.8 (97.9, 99.3) | 96.8 (94.6, 98.3) | 93.4 (91.8, 94.8) | <0.001 |
| **Days post onset<sup>c</sup>** |  |  |  |  |  |  |  |  |  |
| 0 | 15 (20.0) | 55 (73.3) | 0 (0) | 5 (6.7) | 75.0 (50.9, 91.3) | 100 (93.5, 100) | 100 (78.2, 100) | 91.7 (81.6, 97.2) | 0.074 |
| 1–3 | 129 (26.9) | 337 (70.2) | 3 (0.6) | 11 (2.3) | 92.1 (86.4, 96.0) | 99.1 (97.4, 99.8) | 97.7 (93.5, 99.5) | 96.8 (94.4, 98.4) | 0.061 |
| 4–6 | 145 (40.4) | 201 (56.0) | 4 (1.1) | 9 (2.5) | 94.2 (89.2, 97.3) | 98.0 (95.1, 99.5) | 97.3 (93.3, 99.3) | 95.7 (92.0, 98.0) | 0.267 |
| ≥7 | 85 (30.6) | 154 (55.4) | 3 (1.1) | 36 (12.9) | 70.2 (61.3, 78.2) | 98.1 (94.5, 99.6) | 96.6 (90.4, 99.3) | 81.1 (74.7, 86.4) | <0.001 |
| **Symptomatology<sup>d</sup>** |  |  |  |  |  |  |  |  |  |
| Asymptomatic | 37 (11.9) | 252 (81.3) | 3 (1.0) | 18 (5.8) | 67.3 (53.3, 79.3) | 98.8 (96.6, 99.8) | 92.5 (79.6, 98.4) | 93.3 (89.7, 96.0) | 0.002 |
| ≥1 symptom | 353 (29.0) | 797 (65.5) | 10 (0.8) | 56 (4.6) | 86.3 (82.6, 89.5) | 98.8 (97.7, 99.4) | 97.2 (95.0, 98.7) | 93.4 (91.6, 95.0) | <0.001 |
| 1–3 symptoms | 266 (30.5) | 556 (63.8) | 9 (1.0) | 41 (4.7) | 86.6 (82.3, 90.2) | 98.4 (97.0, 99.3) | 96.7 (93.9, 98.5) | 93.1 (90.8, 95.0) | <0.001 |
| 4–6 symptoms | 24 (19.4) | 92 (74.2) | 0 (0) | 8 (6.5) | 75.0 (56.6, 88.5) | 100 (96.1, 100) | 100 (85.8, 100) | 92.0 (84.8, 96.5) | 0.013 |
| 7–9 symptoms | 20 (19.4) | 80 (77.7) | 0 (0) | 3 (2.9) | 87.0 (66.4, 97.2) | 100 (95.5, 100) | 100 (83.2, 100) | 96.4 (89.8, 99.2) | 0.248 |
| ≥10 symptoms | 43 (36.8) | 69 (59.0) | 1 (0.9) | 4 (3.4) | 91.5 (79.6, 97.6) | 98.6 (92.3, 100) | 97.7 (88.0, 99.9) | 94.5 (86.6, 98.5) | 0.371 |
| **Collection strategy** |  |  |  |  |  |  |  |  |  |
| Self, SEDSS | 46 (34.3) | 80 (59.7) | 0 (0) | 8 (6.0) | 85.2 (72.9, 93.4) | 100 (95.5, 100) | 100 (92.3, 100) | 90.9 (82.9, 96.0) | 0.013 |
| Staff, SEDSS | 23 (12.2) | 159 (84.6) | 0 (0) | 6 (3.2) | 79.3 (60.3, 92.0) | 100 (97.7, 100) | 100 (85.2, 100) | 96.4 (92.3, 98.7) | 0.041 |
| Self + Staff, SEDSS | 69 (21.4) | 239 (74.2) | 0 (0) | 14 (4.3) | 83.1 (73.3, 90.5) | 100 (98.5, 100) | 100 (94.8, 100) | 94.5 (90.9, 96.9) | 0.001 |
| Staff, COPA | 322 (26.7) | 813 (67.3) | 13 (1.1) | 60 (5.0) | 84.3 (80.2, 87.8) | 98.4 (97.3, 99.2) | 96.1 (93.5, 97.9) | 93.1 (91.2, 94.7) | <0.001 |
| Staff, SEDSS + COPA | 345 (24.7) | 972 (69.6) | 13 (0.9) | 66 (4.7) | 83.9 (80.0, 87.4) | 98.7 (97.8, 99.3) | 96.4 (93.9, 98.1) | 93.6 (92.0, 95.0) | <0.001 |
| **Repeated tests<sup>e</sup>** |  |  |  |  |  |  |  |  |  |
| Initial test | 105 (57.1) | 72 (39.1) | 3 (1.6) | 4 (2.2) | 96.3 (90.9, 99.0) | 96.0 (88.8, 99.2) | 97.2 (92.1, 99.4) | 94.7 (87.1, 98.5) | 1 |
| Repeated test 7–14 days after initial test | 29 (15.8) | 122 (66.3) | 3 (1.6) | 30 (16.3) | 49.2 (35.9, 62.5) | 97.6 (93.1, 99.5) | 90.6 (75.0, 98.0) | 80.3 (73.0, 86.3) | <0.001 |
| **Number of vaccine doses<sup>f</sup>** |  |  |  |  |  |  |  |  |  |
| Unvaccinated | 99 (19.9) | 378 (75.9) | 0 (0) | 21 (4.2) | 82.5 (74.5, 88.8) | 100 (99.0, 100) | 100 (96.3, 100) | 94.7 (92.1, 96.7) | <0.001 |
| 1 dose | 7 (16.3) | 33 (76.7) | 1 (2.3) | 2 (4.7) | 77.8 (40.0, 97.2) | 97.1 (84.7, 99.9) | 87.5 (47.3, 99.7) | 94.3 (80.8, 99.3) | 1 |
| 2 doses | 100 (22.9) | 310 (71.1) | 2 (0.5) | 24 (5.5) | 80.6 (72.6, 87.2) | 99.4 (97.7, 99.9) | 98.0 (93.1, 99.8) | 92.8 (89.5, 95.3) | <0.001 |
| 3 doses | 144 (38.6) | 201 (53.9) | 8 (2.1) | 20 (5.4) | 87.8 (81.8, 92.4) | 96.2 (92.6, 98.3) | 94.7 (89.9, 97.7) | 91.0 (86.4, 94.4) | 0.038 |
| 4 doses | 2 (22.2) | 6 (66.7) | 1 (11.1) | 0 (0) | 100 (15.8, 100) | 85.7 (42.1, 99.6) | 66.7 (9.4, 99.2) | 100 (54.1, 100) | 1 |
| **Predominant SARS-CoV-2 variant** |  |  |  |  |  |  |  |  |  |
| Pre-Delta | 80 (16.7) | 379 (79.1) | 0 (0) | 20 (4.2) | 80.0 (70.8, 87.3) | 100 (99.0, 100) | 100 (95.5, 100) | 95.0 (92.4, 96.9) | <0.001 |
| Delta | 7 (4.2) | 156 (94.0) | 0 (0) | 3 (1.8) | 70.0 (34.8, 93.3) | 100 (97.7, 100) | 100 (59.0, 100) | 98.1 (94.6, 99.6) | 0.248 |
| Omicron | 304 (34.4) | 517 (58.4) | 13 (1.5) | 51 (5.8) | 85.6 (81.5, 89.1) | 97.5 (95.8, 98.7) | 95.9 (93.1, 97.8) | 91.0 (88.4, 93.2) | <0.001 |

RT-PCR: reverse transcription polymerase chain reaction; CI: confidence interval
<sup>a</sup> There were 1530 tests of both BinaxNOW and RT-PCR.
<sup>b</sup> We used McNemar's test to evaluate differences in proportions of discordant pairs (i.e., the differences between false positives and false negatives) between BinaxNOW and the reference standard, RT-PCR. It helps determine if one test is more likely to produce false positives or false negatives compared to the other.
<sup>c</sup> N = 1192 tests from 923 participants—284 participants were missing symptom onset dates.
<sup>d</sup> N = 1526 tests from 1203 participants—4 participants were missing symptom data. Symptoms included tiredness, cough, loss of smell, dyspnea, myalgia, throat pain, chest pain, nausea/vomiting, diarrhea, abdominal pain, nasal congestion, chills, conjunctivitis, skin changes, rash, arthralgia, eye pain, bleeding, irritability, and calf pain.
<sup>e</sup> N = 368 tests from 184 participants. Restricted to participants who had repeated tests.
<sup>f</sup> N = 1359 tests from 1063 participants—144 participants were missing vaccination data.

The sensitivity of BinaxNOW was higher for symptomatic (86.3%) than for asymptomatic (67.3%) participants, whereas specificity estimates were the same (98.8%) for both groups. Sensitivity did not significantly vary by the number of symptoms reported (Figure S1). For symptomatic participants, one correct diagnosis was obtained for every 1.2 patients tested with BinaxNOW on average during the study period (NND = 1.2, 95% CI: 1.1–1.2) (Table 3). For asymptomatic participants, one correct diagnosis was obtained for every 1.5 patients tested with BinaxNOW on average during the study period (NND = 1.5, 95% CI: 1.3–2.0). The sensitivity and specificity of BinaxNOW showed consistent performance across participants regardless of the number of COVID-19 vaccine doses received, with overlapping confidence intervals for all groups (Table 2).

**Table 3.** Performance of BinaxNOW compared to RT-PCR (N = 1530 paired^a^ tests from 1207 participants unless stated otherwise).

|  | Positive likelihood ratio (95% CI) | Negative likelihood ratio (95% CI) | Correctly classified proportion (95% CI) | Apparent positivity (95% CI) | True positivity (95% CI) | Number needed to diagnose (95% CI) | AUC-ROC |
| --- | --- | --- | --- | --- | --- | --- | --- |
| **Overall** | 68.89 (40.07, 118.41) | 0.16 (0.13, 0.20) | 0.943 (0.930, 0.954) | 0.264 (0.242, 0.287) | 0.304 (0.281, 0.328) | 1.2 (1.2, 1.3) | 0.914 |
| **Days post onset<sup>b</sup>** |  |  |  |  |  |  |  |
| 0 | Inf | 0.25 (0.12, 0.53) | 0.933 (0.851, 0.978) | 0.200 (0.116, 0.308) | 0.267 (0.171, 0.381) | 1.3 (1.1, 2.3) | 0.875 |
| 1–3 | 104.43 (33.81, 322.51) | 0.08 (0.04, 0.14) | 0.971 (0.952, 0.984) | 0.275 (0.236, 0.317) | 0.292 (0.251, 0.335) | 1.1 (1.0, 1.2) | 0.956 |
| 4–6 | 48.25 (18.27, 127.44) | 0.06 (0.03, 0.11) | 0.964 (0.939, 0.981) | 0.415 (0.364, 0.468) | 0.429 (0.377, 0.482) | 1.1 (1.0, 1.2) | 0.961 |
| ≥7 | 36.76 (11.91, 113.43) | 0.30 (0.23, 0.40) | 0.860 (0.813, 0.898) | 0.317 (0.262, 0.375) | 0.435 (0.376, 0.496) | 1.5 (1.3, 1.8) | 0.842 |
| **Symptomatology<sup>c</sup>** |  |  |  |  |  |  |  |
| Asymptomatic | 57.18 (18.29, 178.78) | 0.33 (0.23, 0.48) | 0.932 (0.898, 0.958) | 0.129 (0.094, 0.172) | 0.177 (0.137, 0.225) | 1.5 (1.3, 2.0) | 0.830 |
| ≥1 symptom | 69.65 (37.58, 129.11) | 0.14 (0.11, 0.18) | 0.946 (0.931, 0.958) | 0.299 (0.273, 0.325) | 0.336 (0.310, 0.364) | 1.2 (1.1, 1.2) | 0.925 |
| 1–3 symptoms | 54.39 (28.41, 104.15) | 0.14 (0.10, 0.18) | 0.943 (0.925, 0.957) | 0.315 (0.285, 0.347) | 0.352 (0.320, 0.385) | 1.2 (1.1, 1.3) | 0.925 |
| 4–6 symptoms | Inf | 0.25 (0.14, 0.46) | 0.935 (0.877, 0.972) | 0.194 (0.128, 0.274) | 0.258 (0.184, 0.344) | 1.3 (1.1, 1.9) | 0.875 |
| 7–9 symptoms | Inf | 0.13 (0.05, 0.37) | 0.971 (0.917, 0.994) | 0.194 (0.123, 0.284) | 0.223 (0.147, 0.316) | 1.2 (1.0, 1.6) | 0.935 |
| ≥10 symptoms | 64.04 (9.13, 449.18) | 0.09 (0.03, 0.22) | 0.957 (0.903, 0.986) | 0.376 (0.288, 0.470) | 0.402 (0.312, 0.496) | 1.1 (1.0, 1.4) | 0.950 |
| **Collection strategy** |  |  |  |  |  |  |  |
| Self, SEDSS | Inf | 0.15 (0.08, 0.28) | 0.940 (0.886, 0.974) | 0.343 (0.263, 0.430) | 0.403 (0.319, 0.491) | 1.2 (1.1, 1.5) | 0.926 |
| Staff, SEDSS | Inf | 0.21 (0.10, 0.42) | 0.968 (0.932, 0.988) | 0.122 (0.079, 0.178) | 0.154 (0.106, 0.214) | 1.3 (1.1, 1.7) | 0.897 |
| Self + Staff, SEDSS | Inf | 0.17 (0.10, 0.27) | 0.957 (0.928, 0.976) | 0.214 (0.171, 0.263) | 0.258 (0.211, 0.309) | 1.2 (1.1, 1.4) | 0.916 |
| Staff, COPA | 53.56 (31.18, 92.00) | 0.16 (0.13, 0.20) | 0.940 (0.925, 0.952) | 0.277 (0.252, 0.303) | 0.316 (0.290, 0.343) | 1.2 (1.2, 1.3) | 0.914 |
| Staff, SEDSS + COPA | 63.60 (37.00, 109.32) | 0.16 (0.13, 0.20) | 0.943 (0.930, 0.955) | 0.256 (0.234, 0.280) | 0.294 (0.271, 0.319) | 1.2 (1.2, 1.3) | 0.913 |
| **Repeated tests<sup>d</sup>** |  |  |  |  |  |  |  |
| Initial test | 24.08 (7.94, 73.03) | 0.04 (0.01, 0.10) | 0.962 (0.923, 0.985) | 0.587 (0.512, 0.659) | 0.592 (0.518, 0.664) | 1.1 (1.0, 1.3) | 0.961 |
| Repeated test 7–14 days after initial test | 20.48 (6.50, 64.53) | 0.52 (0.40, 0.67) | 0.821 (0.757, 0.873) | 0.174 (0.122, 0.237) | 0.321 (0.254, 0.393) | 2.1 (1.6, 3.4) | 0.734 |
| **Number of vaccine doses<sup>e</sup>** |  |  |  |  |  |  |  |
| Unvaccinated | Inf | 0.18 (0.12, 0.26) | 0.958 (0.936, 0.974) | 0.199 (0.165, 0.237) | 0.241 (0.204, 0.281) | 1.2 (1.1, 1.4) | 0.913 |
| 1 dose | 26.44 (3.72, 188.16) | 0.23 (0.07, 0.78) | 0.930 (0.809, 0.985) | 0.186 (0.084, 0.334) | 0.209 (0.100, 0.360) | 1.3 (1.0, 4.1) | 0.874 |
| 2 doses | 125.81 (31.52, 502.14) | 0.19 (0.14, 0.28) | 0.940 (0.914, 0.961) | 0.234 (0.195, 0.277) | 0.284 (0.242, 0.329) | 1.2 (1.1, 1.4) | 0.900 |
| 3 doses | 22.94 (11.60, 45.37) | 0.13 (0.08, 0.19) | 0.925 (0.893, 0.950) | 0.408 (0.357, 0.459) | 0.440 (0.389, 0.492) | 1.2 (1.1, 1.3) | 0.920 |
| 4 doses | 7.00 (1.14, 42.97) | 0.00 (0.00, 0.00) | 0.889 (0.518, 0.997) | 0.333 (0.075, 0.701) | 0.222 (0.028, 0.600) | 1.2 (-2.4, 1.0) | 0.929 |
| **Predominant SARS-CoV-2 variant** |  |  |  |  |  |  |  |
| Pre-Delta | Inf | 0.20 (0.14, 0.30) | 0.958 (0.936, 0.974) | 0.167 (0.135, 0.203) | 0.209 (0.173, 0.248) | 1.2 (1.1, 1.4) | 0.900 |
| Delta | Inf | 0.30 (0.12, 0.77) | 0.982 (0.948, 0.996) | 0.042 (0.017, 0.085) | 0.060 (0.029, 0.108) | 1.4 (1.1, 3.1) | 0.850 |
| Omicron | 34.91 (20.37, 59.82) | 0.15 (0.11, 0.19) | 0.928 (0.909, 0.944) | 0.358 (0.327, 0.391) | 0.401 (0.369, 0.434) | 1.2 (1.1, 1.3) | 0.916 |

RT-PCR: reverse transcription polymerase chain reaction; CI: confidence interval; AUC-ROC: Area Under the Receiver Operating Characteristic Curve
<sup>a</sup> There were 1530 tests of both BinaxNOW and RT-PCR.
<sup>b</sup> N = 1192 tests from 923 participants—284 participants were missing symptom onset dates.
<sup>c</sup> N = 1526 tests from 1203 participants—4 participants were missing symptom data. Symptoms included tiredness, cough, loss of smell, dyspnea, myalgia, throat pain, chest pain, nausea/vomiting, diarrhea, abdominal pain, nasal congestion, chills, conjunctivitis, skin changes, rash, arthralgia, eye pain, bleeding, irritability, and calf pain.
<sup>d</sup> N = 368 tests from 184 participants. Restricted to participants who had repeated tests.
<sup>e</sup> N = 1359 tests from 1063 participants—144 participants were missing vaccination data.

We evaluated the diagnostic performance of BinaxNOW using swabs collected and tested by participants, as well as those collected and tested by study staff. BinaxNOW testing of self-collected and staff-collected anterior nasal swabs from SEDSS showed sensitivities of 85.2% and 79.3%, respectively, and 100% specificity (Table 2). BinaxNOW testing of staff-collected anterior nasal swabs from COPA had 84.3% sensitivity and 98.4% specificity. BinaxNOW tests in anterior nasal swabs collected by both participants (AUC-ROC = 0.926) and staff (AUC-ROC = 0.913) showed a strong ability to discriminate between true positives and true negatives (Table 3). Among individuals positive by RT-PCR, SEDSS participants had lower median Ct values (23, IQR: 21–30) compared to symptomatic COPA participants (27, IQR: 23–31) (*p*=0.004) and a higher median number of symptoms (9, IQR: 5–12) compared to symptomatic COPA participants (1, IQR: 1–1) (*p*<0.001). In the COPA cohort, sensitivity was 55.4% (95% CI: 44.1%–66.3%) for 83 positive RT-PCR tests from asymptomatic participants and 86.8% (95% CI: 82.8%–90.1%) for 355 positive RT-PCRs from symptomatic participants (Figure S2).

There were 184 participants who had repeated tests within a single illness or exposure event (7– 14 days after the initial test). In the initial test, BinaxNOW demonstrated high sensitivity (96.3%) and specificity (96.0%) for detecting SARS-CoV-2 (Table 2). During subsequent sample collection and testing 7–14 days later, sensitivity decreased to 48.4%, while specificity remained high at 97.9% (*p*-value from McNemar’s test < 0.001). Restricting to 134 participants who had the initial test within 6 days of symptom onset, the sensitivity was 96.1% for the initial test and 48.8% for the repeated test 7–14 days later. The initial test showed strong overall performance (AUC-ROC = 0.961), whereas the follow-up testing showed a decline in accuracy for identifying positive cases over time (AUC-ROC = 0.731) (Table 3). Sensitivity dropped significantly for participants initially symptomatic (98.7%) and later asymptomatic (23.1%) (Figure 2, Table S2). Conversely, sensitivity increased for those initially asymptomatic (50.0%) and later symptomatic (100%), but this difference was not statistically significant possibly due to the limited sample size.

**Figure 2.**
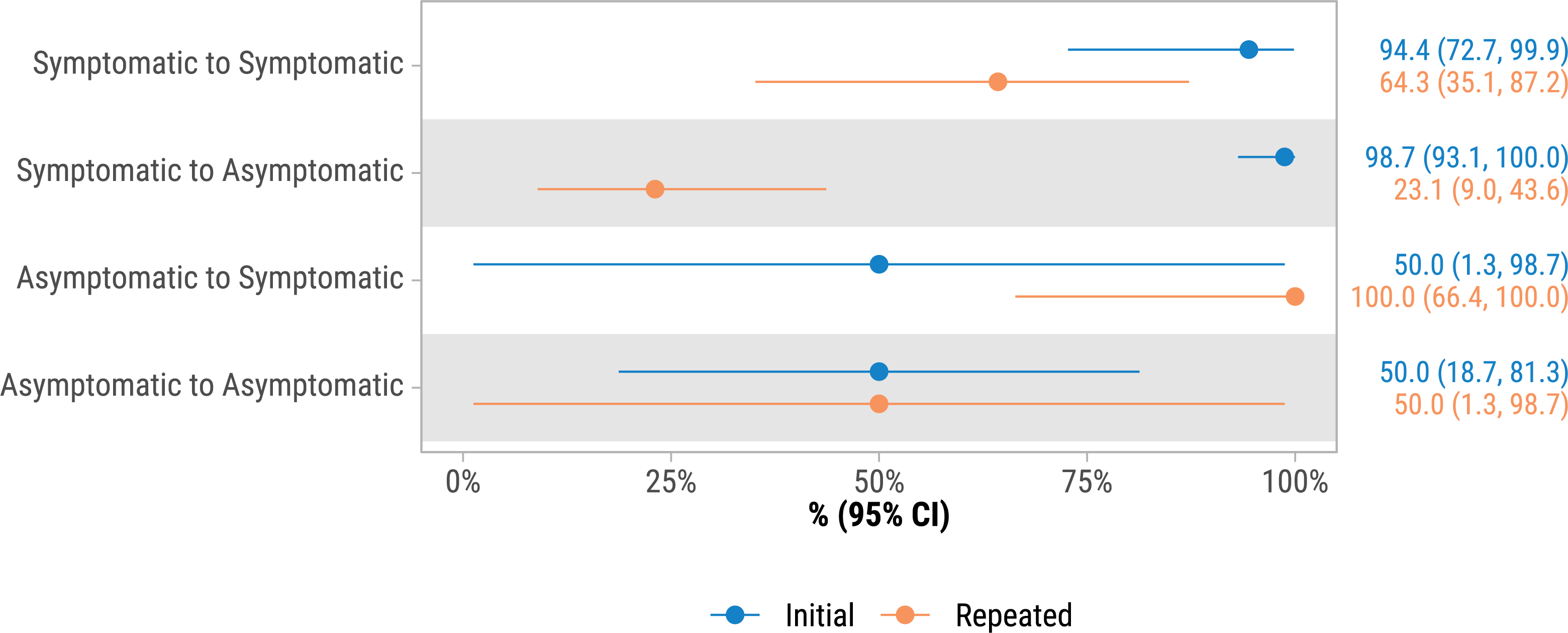
Sensitivity of BinaxNOW Antigen test compared to RT-PCR for initial tests and repeated tests 7–14 days later by symptom status for the initial and repeated tests (N = 368 paired tests from 184 participants). Additional diagnostic accuracy measures are shown in S2 Table. There were 368 tests of both BinaxNOW and RT-PCR.

The sensitivity of BinaxNOW varied significantly depending on the Ct values from positive RT-PCR tests. For Ct values ≤25, paired BinaxNOW tests showed 99.5% sensitivity in correctly identifying positive cases (Table 4). Conversely, as Ct values increased, test accuracy declined, reaching only 18.2% for Ct values between 36–40.

**Table 4.** Sensitivity of BinaxNOW by Ct values of RT-PCR tests (N = 464 paired^a^ tests from 435 participants).

| **Ct value** | **Positive PCR tests**<br>**N** | **Positive BinaxNOW tests**<br>**N (%)** | **Negative BinaxNOW tests**<br>**N (%)** |
| --- | --- | --- | --- |
| ≤25 | 198 | 197 (99.5) | 1 (0.5) |
| 26–30 | 121 | 113 (93.4) | 8 (6.6) |
| 31–35 | 101 | 72 (71.3) | 29 (28.7) |
| 36–40 | 44 | 8 (18.2) | 36 (81.8) |

RT-PCR: reverse transcription polymerase chain reaction.
<sup>a</sup> This table includes positive RT-PCR tests and corresponding BinaxNOW test results. Of 465 total positive RT-PCR tests, 1 was missing a Ct value, therefore the sample size includes 464 tests of both BinaxNOW and RT-PCR.

## Discussion

Our results demonstrated an overall 84.1% sensitivity for the Abbot BinaxNOW COVID-19 Antigen Card Test which falls within the upper range of previously reported BinaxNOW sensitivities (50–85%) among other studies.^11–14,29^ The test also demonstrated high specificity (98.8%), positive predictive value (96.8%), and negative predictive value (93.4%). Test sensitivity was highest 1–6 days post onset and decreased significantly thereafter. These findings are in agreement with other studies, highlighting the importance of timing in SARS-CoV-2 antigen testing.^30,31^

Our findings regarding BinaxNOW test performance in symptomatic and asymptomatic individuals also align with those from other studies,^11–14^ showing substantially higher test sensitivity in symptomatic compared to asymptomatic individuals, while maintaining a high level of specificity for both groups. We did not find a clear dose-response relationship between the number of symptoms experienced and sensitivity, but the point estimate for test sensitivity was highest (91.5%) among participants with ten or more symptoms. Symptom type and indicators of disease severity, such as low oxygen saturation levels, tachypnea, or requiring hospitalization, rather than simply the number of symptoms reported, may have a greater influence on diagnostic accuracy.^32^ These results corroborate previous research and highlight the challenges of detecting SARS-CoV-2 infections in asymptomatic cases.^11–14^ Clinicians should consider these factors and follow CDC guidelines for using antigen tests, including repeat testing for asymptomatic individuals who were exposed, considering other etiologies for symptomatic individuals, and repeating testing with RT-PCR in situations where sensitivity is of paramount importance according to CDC recommendations.^33^

Following infection, SARS-CoV-2 viral replication and shedding precede symptoms, with peak viral titers occurring near the day of symptom onset and declining thereafter.^34^ This trend is supported by studies indicating that antigen testing demonstrates higher sensitivity early in infection when viral loads are high, while repeated sampling over the illness course correlates with decreasing sensitivity.^11,30,31,35,36^ Ct values from RT-PCR tests also provide quantity of viral genetic material in the sample (as an approximate proxy for viral load) with increasing Ct values reflecting decreasing viral genetic material.^37,38^ Our study used the same RT-PCR assay for SEDSS participants, but two different RT-PCR assays were used in COPA, which precludes direct comparison of Ct values due to variation in sensitivity, chemistry of reagents, gene targets, cycle parameters, and others.^37^ BinaxNOW test showed peak sensitivity (99.5%) when the Ct values of paired RT-PCR tests were 25 or lower, suggesting a higher concentration of viral genetic material, typically indicative of early-stage infection. This is consistent with our findings of reduced sensitivity 7 or more days after symptom onset, as well as those showing a significant decline in sensitivity with repeated testing conducted in samples collected 7–14 days after initial testing. These findings emphasize the importance of testing during the early infection stage and maximizing the utility of isolation and treatment, when indicated. However, BinaxNOW test sensitivity drops significantly (18.2%) for cases with Ct values between 36–40, suggesting a diminished capacity to detect positive SARS-CoV-2 cases among individuals with lower viral genetic material concentrations during later stages of infection.

Compared to ancestral variants, Delta and Omicron are characterized by their shorter incubation periods, serial intervals, enhanced immune evasion, and heightened transmissibility.^39–41^ Studies have yielded mixed results in viral load patterns for these variants, with some reporting higher viral loads for Delta,^42,43^ whereas others report higher viral loads for Omicron BA.1.^44,45^ The limited number of tests during the Delta variant dominant period in our study precluded robust comparisons of sensitivity between SARS-CoV-2 variants, and there were overlapping confidence intervals for sensitivity across the variants. One study reported lower BinaxNOW COVID-19 Antigen test sensitivity for infections with the Omicron variant compared to those with the Delta variant,^46^ and another found no significant difference in sensitivity between the two variants.^47^ The impact of infection prevalence, such as the lower prevalence in the Delta period, may have affected the results. Lower prevalence can lead to higher false-negative rates as the proportion of true negatives in the population increases, influencing the balance of sensitivity and specificity. Sensitivity and specificity of the BinaxNOW test remained consistent across participants regardless of their COVID-19 vaccination status, similar to other studies.^29,48^

Test timing, the patient’s clinical presentation, and the prevalence of SARS-CoV-2 infection in the community should be considered when interpreting results and making diagnostic decisions. This approach aligns with CDC guidance on COVID-19 testing.^49^ Different settings require tailored testing strategies. Healthcare settings attending to immunocompromised individuals may rely on highly sensitive RT-PCR tests to accurately detect prolonged viral shedding. Conversely, antigen tests may provide sufficient diagnostic accuracy in most settings, particularly when timely results are essential for public health intervention or treatment.

Our study evaluated the performance of BinaxNOW COVID-19 Antigen test for both self-collected and staff-collected anterior nasal swab samples. We observed high sensitivities (85.2% and 83.9%) and specificities (>98%) for both collection methods, consistent with the literature emphasizing the feasibility and reliability of self-collection methods.^50,51^ Sensitivity among participants from the hospital-based surveillance site (SEDSS) (83.1%) was not significantly different than among symptomatic participants from the community-based cohort (COPA) (86.8%), but was significantly higher than asymptomatic COPA participants (55.4%).

This study had several limitations. Our population was composed primarily of individuals that identified as Hispanic/Latino and Puerto Rican between the ages of 0 and 50 years, which may not fully represent diverse populations or epidemiological conditions found elsewhere. BinaxNOW performance may vary in populations with different demographic characteristics, vaccination rates, or healthcare access. Additionally, participants in our study, comprising individuals seeking medical attention or enrolling in a community-based cohort study, may differ from non-participants regarding healthcare-seeking behavior, symptom severity, proximity to healthcare facilities, access to healthcare, socioeconomic factors, and risk perception, potentially introducing selection bias. Our study included pre-Delta, Delta, and Omicron (time period covering BA.1 through XBB.1.5^52^) variants. However, our study population had low SARS-CoV-2 transmission prior to the Omicron variant. More recent Omicron subvariants like EG.5 and FL.1.5.1 may have viral mutations that affect BinaxNOW performance.

Furthermore, our study used the dominant variant period as a proxy for the actual variant of the individual, potentially misclassifying cases due to variability within these periods. Lastly, our study focused on a rapid antigen test for SARS-CoV-2 from a single manufacturer. Our findings may not apply to other antigen tests with potentially different performance characteristics.

## Conclusions

Our study provides valuable insights into the diagnostic performance of BinaxNOW COVID-19 Antigen Card Test in different epidemiological contexts. While demonstrating high sensitivity and specificity, our findings highlight the influence of factors such as symptomatology, viral load, and timing of specimen collection on test accuracy. BinaxNOW remains a valuable tool for home use and early infection identification, offering numerous advantages, including low cost, extended shelf life, temperature stability, ease of use, and the ability to identify individuals with high viral loads. However, its application should be considered alongside clinical and epidemiological context.^33^ Future research should continue to explore the evolving landscape of SARS-CoV-2 variants and the performance of rapid antigen tests across diverse populations to further enhance our understanding and response to COVID-19.

## Data availability

 Due to data security and confidentiality guidelines, all analyses, and restricted-use datasets, including questionnaire forms and code, must be requested from CDC and PMFS after submitting a concept proposal to.

## Ethics approval and consent to participate

Approval for the COPA project was obtained from the Ponce Medical School Foundation, Inc. Institutional Review Board (protocol number 171110-VR). The Institutional Review Boards at the CDC, Auxilio Mutuo, and Ponce Medical School Foundation approved the SEDSS study protocols 6214, and 120308-VR, respectively. Written consent to participate was obtained from all adult participants and emancipated minors; parental written consent and participant assent were obtained for children.

## Disclaimer

The findings and conclusions in this report are those of the authors and do not necessarily represent the official position of the US Centers for Disease Control and Prevention.

