## Supplement for "Diagnostic Accuracy of the Abbot BinaxNOW COVID-19 Antigen Card Test, Puerto Rico"

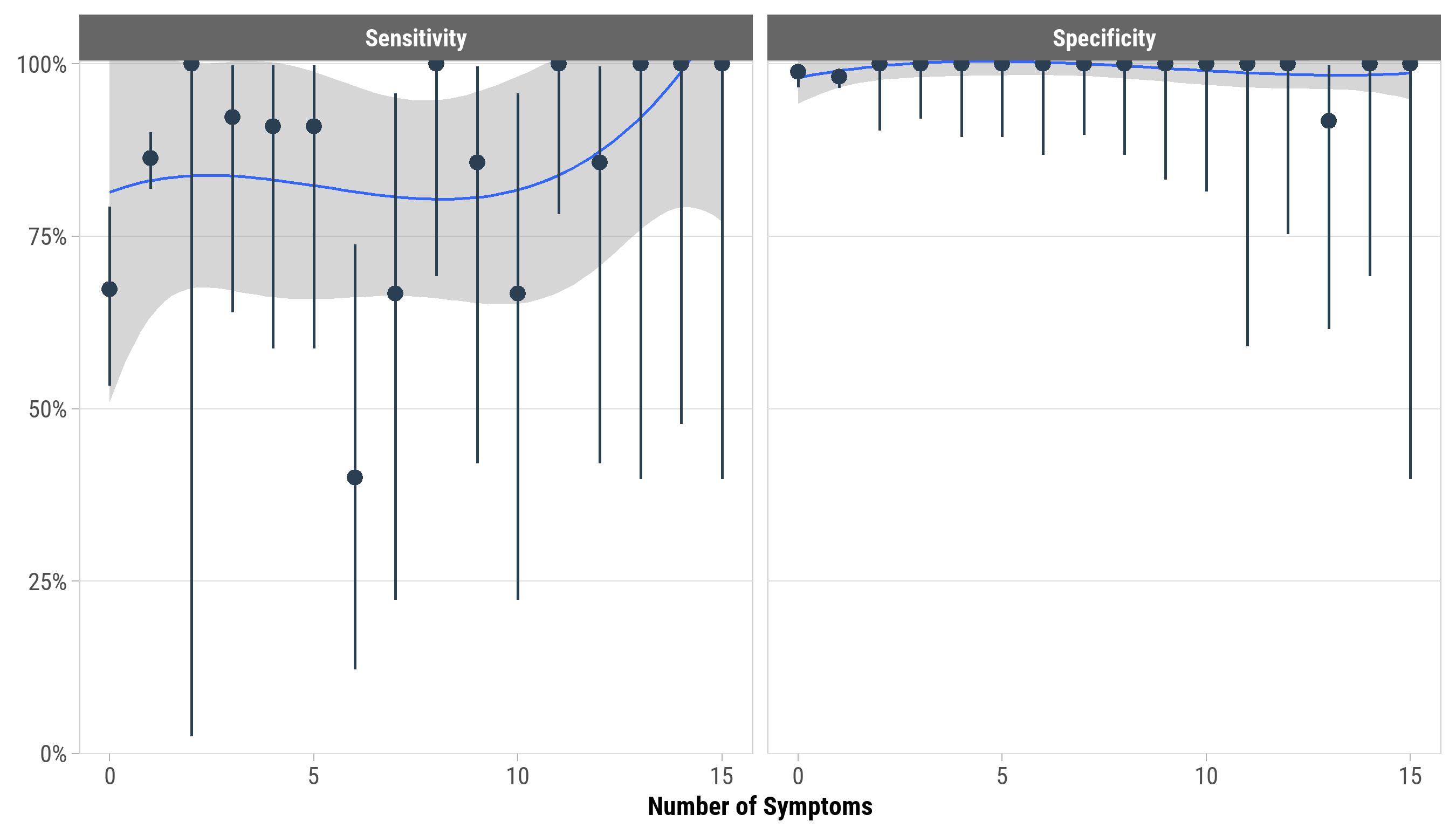

**Figure S1. Sensitivity and specificity of BinaxNOW Antigen test compared to RT-PCR by number of COVID-19 symptoms (N = 1524 paired tests from 1201 participants experiencing 0 to 16 symptoms).** The blue line represents a cubic spline and grey bands are 95% confidence intervals of the model fit. Vertical bars are 95% confidence intervals of the BinaxNOW sensitivity and specificity for each number-of-symptoms subgroup. Symptoms included tiredness, cough, loss of smell, dyspnea, myalgia, throat pain, chest pain, nausea/vomiting, diarrhea, abdominal pain, nasal congestion, chills, conjunctivitis, skin changes, rash, arthralgia, eye pain, bleeding, irritability, and calf pain. There were 1524 tests of both BinaxNOW and RT-PCR.

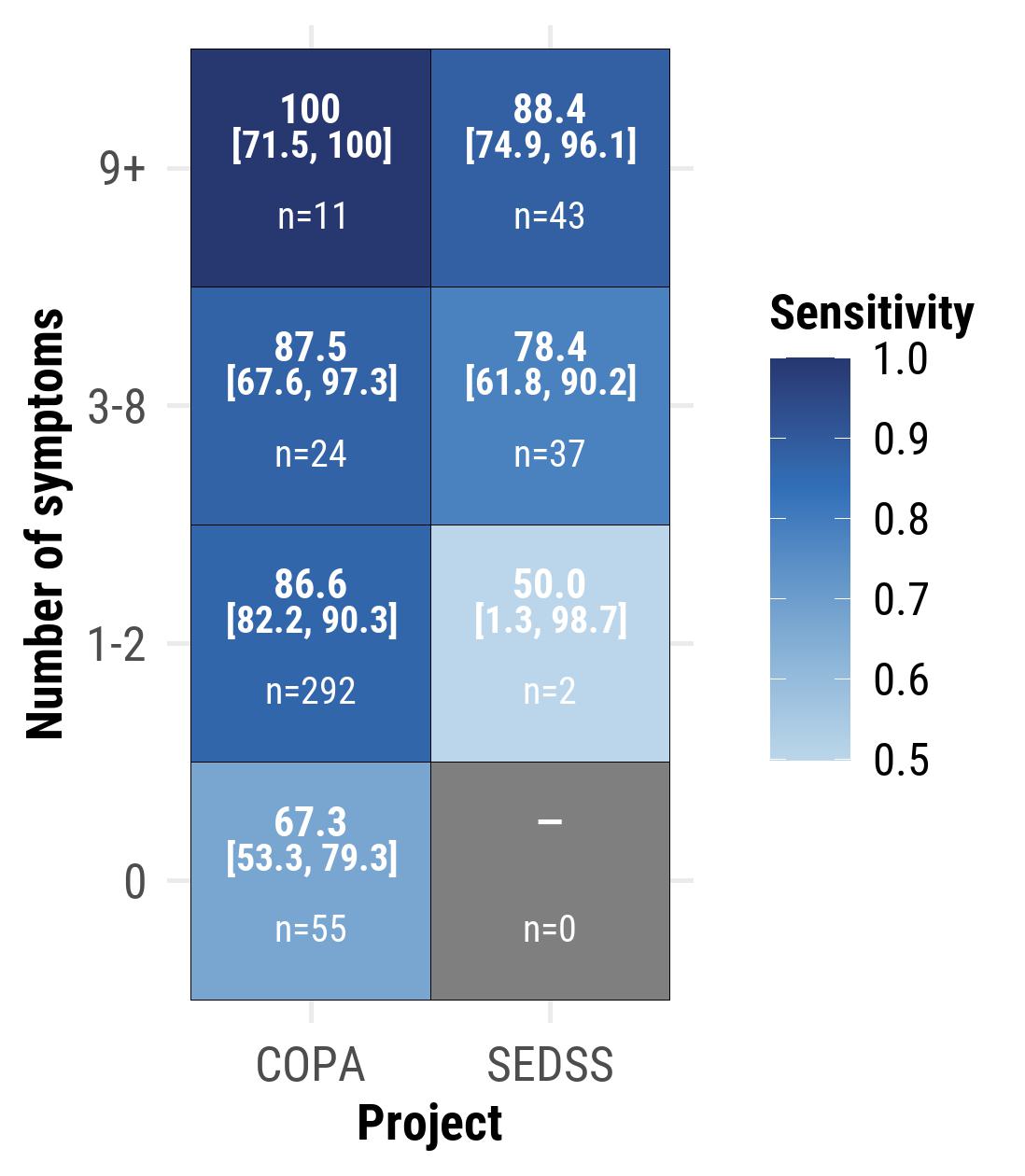

**Figure S2. Sensitivity of BinaxNOW Antigen test compared to RT-PCR by project (COPA, SEDSS) and number of symptoms reported (N = 1526 paired tests from 1203 participants).** Sensitivity, 95% confidence intervals, and the number of positive RT-PCR tests for each group are shown. There were 1526 tests of both BinaxNOW and RT-PCR. Symptoms included tiredness, cough, loss of smell, dyspnea, myalgia, throat pain, chest pain, nausea/vomiting, diarrhea, abdominal pain, nasal congestion, chills, conjunctivitis, skin changes, rash, arthralgia, eye pain, bleeding, irritability, and calf pain.

| Table S1. Definitions of Diagnostic Performance Metrics | |
| --- | --- |
| Term | **Definition** |
| Sensitivity | The proportion of true positive BinaxNOW tests (correctly identified infections) compared to RT-PCR as the reference standard. |
| Specificity | The proportion of true negative BinaxNOW tests (correctly identified non-infections) compared to RT-PCR as the reference standard. |
| Positive predictive value | The probability that a positive BinaxNOW result correctly indicates an actual SARS-CoV-2 infection, as determined by RT-PCR. |
| Negative predictive value | The probability that a negative BinaxNOW result correctly indicates the absence of a SARS-CoV-2 infection, as determined by RT-PCR. |
| Positive likelihood ratio | The likelihood of obtaining a positive BinaxNOW result for SARS-CoV-2 infection compared to RT-PCR, indicating diagnostic performance. |
| Negative likelihood ratio | The likelihood of obtaining a negative BinaxNOW result for SARS-CoV-2 infection compared to RT-PCR, indicating diagnostic performance. |
| Correctly classified proportion | The proportion of participants correctly classified as either SARS-CoV-2 positive or negative by BinaxNOW compared to RT-PCR results. |
| Apparent positivity | The proportion of positive BinaxNOW results in the study population, which includes both correct and incorrect diagnoses by the test. |
| True positivity | The actual proportion of SARS-CoV-2 positive individuals, determined by RT-PCR, regardless of BinaxNOW results. |
| Number needed to diagnose | The number of patients who need to be tested with BinaxNOW to correctly diagnose one person with a SARS-CoV-2 infection, as determined by RT-PCR, in the study population. |

| Table S2. Comparison of BinaxNOW and RT-PCR for initial tests and repeated tests 7–14 days later by symptom status for the initial and repeated tests (N = 368 paired^a^ tests from 184 participants). | | | | | | | | | |
| --- | --- | --- | --- | --- | --- | --- | --- | --- | --- |
|  | **True Positive,**  **n (%)** | **True Negative,**  **n (%)** | **False Positive,**  **n (%)** | **False Negative,**  **n (%)** | **Sensitivity**  **% (95% CI)** | **Specificity**  **% (95% CI)** | **Positive Predictive Value**  **% (95% CI)** | **Negative Predictive Value**  **% (95% CI)** | **McNemar’s Chi Square *P* value** |
| Symptomatic to Symptomatic | | | | | | | | | |
| First | 17 (37.8) | 26 (57.8) | 1 (2.2) | 1 (2.2) | 94.4 (72.7, 99.9) | 96.3 (81.0, 99.9) | 94.4 (72.7, 99.9) | 96.3 (81.0, 99.9) | 1 |
| Second | 9 (20.0) | 31 (68.9) | 0 (0) | 5 (11.1) | 64.3 (35.1, 87.2) | 100 (88.8, 100) | 100 (66.4, 100) | 86.1 (70.5, 95.3) | 0.074 |
| Symptomatic to Asymptomatic | | | | | | | | | |
| First | 78 (85.7) | 10 (11.0) | 2 (2.2) | 1 (1.1) | 98.7 (93.1, 100) | 83.3 (51.6, 97.9) | 97.5 (91.3, 99.7) | 90.9 (58.7, 99.8) | 1 |
| Second | 6 (6.6) | 63 (69.2) | 2 (2.2) | 20 (22.0) | 23.1 (9.0, 43.6) | 96.9 (89.3, 99.6) | 75.0 (34.9, 96.8) | 75.9 (65.3, 84.6) | <0.001 |
| Asymptomatic to Symptomatic | | | | | | | | | |
| First | 1 (5.6) | 16 (88.9) | 0 (0) | 1 (5.6) | 50.0 (1.3, 98.7) | 100 (79.4, 100) | 100 (2.5, 100) | 94.1 (71.3, 99.9) | 1 |
| Second | 9 (50.0) | 9 (50.0) | 0 (0) | 0 (0) | 100 (66.4, 100) | 100 (66.4, 100.0) | 100 (66.4, 100) | 100 (66.4, 100) | 1 |
| Asymptomatic to Asymptomatic | | | | | | | | | |
| First | 5 (16.7) | 19 (63.3) | 1 (3.3) | 5 (16.7) | 50.0 (18.7, 81.3) | 95.0 (75.1, 99.9) | 83.3 (35.9, 99.6) | 79.2 (57.8, 92.9) | 0.221 |
| Second | 1 (5.6) | 16 (88.9) | 0 (0) | 1 (5.6) | 50.0 (1.3, 98.7) | 100 (79.4, 100) | 100 (2.5, 100) | 94.1 (71.3, 99.9) | 1 |
| RT-PCR: reverse transcription polymerase chain reaction; CI: confidence interval  ^a^ There were 368 tests of both BinaxNOW and RT-PCR. | | | | | | | | | |
